# Computer Vision Scoring of Figure Copy and Recall

**DOI:** 10.64898/2026.06.10.26355298

**Authors:** David L. Woods, Kathleen Hall, Isabella Jaramillo, Mike Blank, Kristin Geraci, Andrew Boghossian, Peter Pebler

## Abstract

**Objective:** Figure copy and recall tests are sensitive measures of visuoconstruction and visual episodic memory, but their clinical is constrained by labor-intensive manual scoring. We developed and validated an automated, element-level scoring pipeline using Vertex AI object detection for the tablet-based figure copy and recall tasks in the California Cognitive Assessment Battery (CCAB). The automated scoring pipeline duplicated the scoring procedures used by expert manual raters.

**Methods:** A normative sample of 2,011 community-dwelling adults aged 18-90 completed figure copy and delayed recall trials at baseline, with subsamples retested at 1 day and at 6, 18, and 30 months. Participants completed the drawings with their index finger on a tablet computer with finger position digitized to analyze the speed and timing of individual drawing strokes A convolutional object-detection model trained on the Vertex AI AutoML Vision platform identified each of twelve canonical figure elements in rendered drawings. Separate element presence and location scores were computed after homographically warping drawings onto a canonical template to produce trial-level Element, Location, and Total scores. To compare Vertex and human scores, Vertex AI and expert human raters independently scored 1500 randomly selected drawings to evaluate inter-rater agreement, including a common subset of 100 drawings scored by Vertex AI and all raters.

**Results:** Total scores were virtually indistinguishable (r = 0.966) from human-human agreement (mean r = 0.971) as were Element presence scores (mean r = 0.959 vs. r = 0.963). Location-score agreement (r = 0.951) was slightly below the human-human mean (r = 0.972) due to pixel-level analysis by Vertex AI that was impossible for human raters. The Vertex pipeline showed no preferential advantage for the single expert rater who categorized Elements during training. Automated scores showed strong demographic gradients, age effects on Recall (r = -0.32) were approximately twice those in Copy conditions (r = -0.16). A Memory Cost score (Recall - Copy) showed a monotonic age-related decline from +0.40 z in the youngest subjects to -0.54 z in the oldest. Kinetic analysis revealed that drawing speed and efficiency showed significant age-related changes. Overnight test-retest reliability was high (Recall r = 0.72) and the Recall trial showed a large overnight learning effect (&[Delta] = +1.18) that continued with repeated tests up to 30 months (&[Delta] = +0.75).

**Conclusions:** The computer vision pipeline described here preserves the element-level structure of expert manual scoring and recovers the memory-specific clinical signals of the delayed recall trial. Overall agreement with human rates was approximately an order of magnitude greater than previously described automated scoring approaches. Automatic, computer vision scoring of figure copy and delayed recall removes scoring barriers that impede the more extensive use of figure copy and recall tests.

**Key Findings:**

- A fully automated computer vision pipeline incorporated Vertex AI to score element presence and location in a figure copy and recall task in 9,117 drawings from 2,011 cognitively normal adults,
- In validation samples, score agreement between Vertex AI score and three expert human raters was indistinguishable from human–human agreement (r = 0.966 vs. r = 0.971).
- Age effects on delayed Recall scores were twice those of Copy scores (r = −0.32 vs. r = −0.16). As a result, a Memory Cost score (Recall − Copy in z units) declined monotonically with age from +0.40 z in the youngest stratum to −0.54 z in the oldest.
- Test–retest reliability was high at a 1-day retest interval for Recall (r = 0.72) but obscured by ceiling effects for Copy (r = 0.48). The Recall trial showed large overnight learning effect (Δz = +1.18) that persisted up to 30 months (Δz = +0.75).
- A parallel motor-temporal analysis of drawing time, stroke count, and pause structure revealed that older subjects produced jerkier strokes (r = 0.19) and showed longer intersegment pauses during recall (r = 0.19).

## Introduction

Figure copy and recall tests have been a cornerstone of clinical neuropsychology for more than eight decades. The Rey-Osterrieth Complex Figure (Rey-O), introduced by Rey in 1941 and operationalized by Osterrieth in 1944, and the Benson Complex Figure Test (BCFT), developed for the National Alzheimer’s Coordinating Center Uniform Data Set (UDS) share a common structure: the patient first copies a complex geometric figure with the stimulus in view, then reproduces it from memory after a delay (typically 20–30 minutes). The copy trial probes visuospatial perception, constructional planning, executive organization, and fine motor control. The recall trial probes encoding, consolidation, and retrieval of complex non-verbal visual information. Because figure drawing engages many cognitive systems, including visuoconstruction and visual episodic memory, figure drawing tests are sensitive to a wide range of neurodegenerative conditions and remain a fixture of dementia evaluation, traumatic brain injury assessment, and longitudinal aging research.

However, despite the centrality of these tests, their administration and scoring have changed little since the 1940s. The Rey-O and BCFT are scored using element-based rubrics in which each major component of the target figure is judged on multiple semi-independent criteria. The Osterrieth 18-item system for the Rey-O assigns 0, 0.5, 1, or 2 points to each element based on accuracy and placement (Osterrieth, 1944). The BCFT includes eight major elements, each scored with one point for element shape and one for placement, plus a bonus point for an overall well-formed figure (maximum 17; Possin et al., 2011; Kramer, 2011). Crucially, identical scoring rules are applied on the recall trial, where elements are routinely distorted, fragmented, or displaced. Recall scoring is therefore where rater uncertainty is greatest and where item-level disagreement is most consequential.

The published reliability literature on these scoring systems is, on its surface, reassuring. For the Rey-O, Liberman and colleagues (1994) reported inter-rater correlation coefficients of 0.88 and 0.96 for copy, and delayed recall total scores, with intra-rater coefficients of 0.96–0.99. Tupler and colleagues (1995), studying memory-impaired older adults, reported inter- and intra-rater coefficients of 0.85–0.97 for total scores while the Boston Qualitative Scoring System reports inter-rater reliability of 0.90 or higher for five of its six summary scales (Stern et al., 1999). For the BCFT figure, the original FTLD training materials report an intraclass correlation of 0.95 across 14 raters scoring 10 figures (Kramer, 2011). However, these summary statistics conceal several important problems.

First, total score reliability masks substantial item-level disagreement. In the Tupler et al. study, with total-score reliabilities ranging from 0.85 to 0.97, reliabilities for the 18 individual items ranged from 0.14 to 0.96. While item-level disagreement averaged out across the 18-item test, reliable item-level scores are needed for qualitative analyses, error pattern profiling, and process-based interpretation. Recall scores, with some elements partially drawn or mislocated, is the most clinically informative and the least reliable. Even in studies that report explicit, criterion-anchored scoring rules (Duley et al., 1993) where total scores of independent reviewers falling within two points of each other on roughly 90% of copies and 88% of recalls, maximum disagreements of five points was not uncommon and tended to impact mid-range scores whose classification status is most uncertain.

Second, published reliability estimates were obtained under conditions far more favorable than those encountered in routine clinical practice. Reliability studies use trained raters working from a shared, recently reviewed scoring manual, scoring a manageable batch of figures in a focused session, often after explicit calibration. Clinical practice and multi-site studies involves raters with different familiarity with scoring rules, with rater drift, fatigue, and idiosyncratic accommodation of ambiguous cases. The gap between published training-condition reliability and field reliability is likely substantial but has not been measured directly.

Indeed, even identical raters may score the same figure differently on two occasions. For example, Tupler and colleagues had three raters re-score figures after a three-month interval and reported intra-rater coefficients of 0.85–0.97 for total scores; Liberman et al. reported intra-rater correlations of 0.96–0.99. These coefficients are reassuring at the total-score level but, like the inter-rater estimates, likely mask considerable item-level disagreement.

Another fundamental limitation of manually administered tests is that they only capture the static endpoint of the drawing process and discard information about drawing kinetics and the ordering of elements (Ruffolo et al., 2001). Stroke speed, the timing and number of pauses, intra-stroke smoothness, the order in which elements are produced, and the degree to which a subject draws major structural elements before details, carry useful diagnostic information that cannot be easily recovered from the finished product. Kim and colleagues (2020), using a digital pen and tablet, demonstrated that patients with both early- and late-onset Alzheimer disease copy the Rey-O in a more fragmented manner, with longer inter-stroke pauses, than cognitively normal controls. These differences remain invisible to manual scoring of the finished figure. Process-level abnormalities have also been documented in essential tremor (Peters et al., 2021) and subcortical vascular cognitive impairment (Kim et al., 2025). When only the static drawing is scored, this information is lost from the clinical record.

While figure drawing tests are sensitive, laborious scoring leads to underutilization in neuropsychological practice. The Rey-O is widely regarded as scoring-intensive and is often deferred to specialist evaluations or research protocols. A clinician who must score three Rey-O trials, at up to 10–15 minutes each, is making a meaningful time investment per patient, and the marginal value of that investment is diminished by the rater’s own scoring uncertainty. Even the simplified BCFT, with its eight-element design, requires 3-5 minutes to score each test, an up to 10 minutes with careful checking of borderline elements. As a result, figure drawing and recall tests are deployed less often than their sensitivity warrants.

Similar scoring challenges arise in tests that require subjects to reproduce figures from memory. The Wechsler Memory Scale Visual Reproduction subtest, in continuous use in some form since 1945, presents geometric designs for 10 seconds each and asks the patient to draw each one immediately after presentation, with a delayed reproduction trial after 20–30 minutes (Wechsler, 2009). The Benton Visual Retention Test (BVRT), originally published in 1945 and now in its fifth edition (Benton, 1945; Sivan, 1992), presents ten designs sequentially and asks for immediate reproduction after each, with optional delay administrations and alternate forms for repeated testing. Visual Reproduction and the BVRT are powerful and well-validated measures of visual episodic memory, and in factor-analytic work, the delayed reproduction trial of Visual Reproduction loads primarily on memory rather than on visuospatial intelligence (Larrabee, Kane, Schuck, & Francis, 1985) which aligns it with the Rey-O and Benson recall trials.

### Automated Figure Drawing scoring

Advances over the past decade have demonstrated the technical feasibility of automated scoring. The published approaches fall into three architectural classes, which differ both in performance and in their interpretability to a clinician trained in the underlying scoring system.

The first and most active class is end-to-end deep learning, in which a convolutional or transformer network ingests an image of the finished drawing and emits a single total score. Vogt and colleagues (2019) trained a deep network to score the Rey-O directly from images and reported r = 0.88 with human raters. Park and colleagues (2023) trained a DenseNet on 20,040 drawings from the Gwangju Alzheimer’s and Related Dementia cohort and reported small absolute errors against a five-expert gold standard. Langer and colleagues (2024), in a substantially larger eLife study using more than 20,000 hand-drawn Rey-O images and crowdsourced human ratings, demonstrated that end-to-end convolutional neural network (CNN) scoring is robust to rotation, brightness, and contrast variation, and moreover outperformed clinicians on absolute-error metrics. A public benchmark dataset has since been released to enable comparison across architectures (Guerrero-Martín et al., 2024). A recent review by Andersen, Lundervold, and Ronold (2026) identified five published end-to-end and rule-based systems for the Rey-O trained on more than 41,000 figures with reported agreement against expert raters in the same range as the inter-rater agreement among experts themselves. These end-to-end systems are accurate and consistent, but they have a fundamental limitation: the score is a total score only leaving item level scores inaccessible.

The second class consists of hybrid feature-engineering systems, of which the DCTclock developed at MIT and the Lahey Clinic is the most fully developed and the most widely deployed (Souillard-Mandar et al., 2021). The DCTclock, now an FDA-listed Class II medical device commercialized by Linus Health, captures clock drawings on a digitizing pen or tablet and extracts more than 700 low-level features from the time-stamped pen-stroke record, aggregated by Lasso-regularized logistic regression into eight functional composite scales. The DCTclock has been shown to detect amyloid-positive cognitively unimpaired individuals (Rentz et al., 2021), to distinguish Parkinson disease phenotypes, and to achieve good test-retest reliability (REF). However, while the eight composites have domain labels, a clinician cannot inspect a DCTclock report and verify, as they would for manually scored drawings, that the patient drew each canonical element correctly. The system provides a more transparent intermediate layer than an end-to-end CNN, but element-level scores are still missing.

A new architectural class, introduced here, uses computer vision to mirror manual element-level decisions directly. In this approach, an object detection model trained on annotated figure drawings identifies each element in a drawing, homographically warped into template space, with an element label, a bounding box, and a confidence score. Element presence is scored based the confidence score, and location is scored by comparing of detected bounding box coordinates with those of the template for the identified element. Thus, the model incorporates the accuracy and location criteria identical to those used by human raters. The total score is the sum of element presence and location scores as in Osterrieth and Kramer scoring methods. This architecture preserves what end-to-end and hybrid systems discard: every scoring decision is exposed at the element level, visualized as a labeled bounding box on the drawing, and can be compared to manually scored data.

This paper describes a transparent, element-level automated scoring pipeline for figure copy and recall tests, developed and validated on a normative sample of 2,011 adults aged 18–90. Drawings were completed on touchscreen tablet computers during normative data collection for the California Cognitive Assessment Battery (Woods et al., 2024). For each drawing, a Vertex AI object detection model trained on 1,774 manually scored figures identified each of twelve canonical element types and assigned a corresponding bounding box and a confidence score. Element-level presence and location total scores were then computed. We report five sets of analyses: (1) agreement of the automated pipeline with three expert human raters, benchmarked against human–human agreement; (2) the demographic and clinical correlates of the automated scores in the full normative sample; (3) age-related decline on the recall trial and on a Memory Cost difference score; (4) test– retest reliability and overnight retest learning across 1-day through 30-month retest waves; (5) results from complementary motor-temporal pipeline characterizing stroke-level timing features from the time-stamped fingertip position log.

Automated figure scoring need not be a black box. By automating the classical scoring method and exposing scoring decisions for review, computer-vision scoring transforms figure copy and recall tests from a laborious process with subjective measures into fully automated, efficient, and auditable process yielding objective measures of visuoconstruction and visual episodic memory.

## Methods

### Participants

Participants were 2,011 community-dwelling adults drawn from a diverse normative sample who completed the California Cognitive Assessment Battery (CCAB; Woods et al., 2024), a tablet-administered cognitive battery developed by Neurobehavioral Systems for clinical and research use across the adult lifespan. The sample comprised subjects who completed the Copy and Recall trials of the figure drawing task at enrollment, with telemedical proctored test administration in their homes. Inclusion required age ≥ 18 years, adequate corrected vision and hearing for tablet-based testing, and absence of current major neurological diagnosis (stroke, dementia, Parkinson disease) or active psychiatric crisis at screening. Recruitment targeted demographic balance across age, sex, education, and racial/ethnic background. All participants completed informed consent under a protocol approved by Western Institutional Review Board. Demographic and clinical measures collected at enrollment included years of education, the CCAB vocabulary subtest, the Cognitive Failures Questionnaire (CFQ), the Geriatric Depression Scale (GDS), the Generalized Anxiety Disorder scale (GAD), self-reported daily medication count, self-reported daily computer use and reading hours, and the FS20 functional status measure. Demographic characteristics of the participants by age quintile are summarized in Table 1.

**Table 1.** Demographic and clinical characteristics of the analytic sample by age quintile. Values are mean ± SD unless otherwise indicated. CFQ = Cognitive Failures Questionnaire; GDS = Geriatric Depression Scale; GAD-7 = Generalized Anxiety Disorder 7-item scale; FS20 = 20-item functional status measure. Vocabulary is the CCAB QSIN vocabulary score.

| Variable | 18–30 | 31–45 | 46–60 | 61–75 | 76–90 | Total |
| --- | --- | --- | --- | --- | --- | --- |
| **N** | 274 | 422 | 542 | 623 | 150 | 2011 |
| **Age (years)** | 25.3 ± 3.3 | 37.3 ± 4.2 | 53.9 ± 4.1 | 67.7 ± 4.2 | 79.3 ± 3.4 | 52.7 ± 17.1 |
| **Sex, n female (% F)** | 160 (60%) | 225 (55%) | 306 (57%) | 375 (60%) | 66 (44%) | 1132 (57%) |
| **White, n (%)** | 64 (23%) | 117 (28%) | 191 (35%) | 280 (45%) | 85 (57%) | 737 (37%) |
| **Asian, n (%)** | 64 (23%) | 77 (18%) | 84 (15%) | 105 (17%) | 12 (8%) | 342 (17%) |
| **Black, n (%)** | 41 (15%) | 71 (17%) | 114 (21%) | 150 (24%) | 36 (24%) | 412 (20%) |
| **Other, n (%)** | 105 (38%) | 157 (37%) | 153 (28%) | 87 (14%) | 16 (11%) | 518 (26%) |
| **Education (years)** | 14.8 ± 2.1 | 15.3 ± 2.2 | 15.3 ± 2.2 | 16.0 ± 2.4 | 16.4 ± 2.5 | 15.5 ± 2.3 |
| **Vocabulary** | 29.9 ± 9.3 | 32.6 ± 8.1 | 32.8 ± 8.3 | 36.9 ± 9.9 | 39.1 ± 9.0 | 34.1 ± 9.4 |
| **Daily computer use (hrs)** | 4.5 ± 2.2 | 4.9 ± 2.1 | 4.5 ± 2.3 | 4.2 ± 1.9 | 3.9 ± 1.7 | 4.4 ± 2.1 |
| **Daily reading (hrs)** | 3.5 ± 3.5 | 4.8 ± 4.2 | 4.4 ± 4.1 | 5.9 ± 4.7 | 6.7 ± 4.7 | 5.0 ± 4.4 |
| **Comorbidity count** | 0.7 ± 1.0 | 0.9 ± 1.2 | 1.2 ± 1.5 | 1.6 ± 1.7 | 1.9 ± 1.7 | 1.3 ± 1.5 |
| **Daily medications** | 0.6 ± 1.2 | 0.7 ± 1.3 | 1.5 ± 2.1 | 2.4 ± 2.1 | 2.9 ± 2.0 | 1.6 ± 2.0 |
| **CFQ score** | 36.9 ± 14.5 | 33.9 ± 14.9 | 30.9 ± 15.9 | 28.8 ± 13.3 | 28.8 ± 12.6 | 31.5 ± 14.8 |
| GDS score | 4.6 ± 3.7 | 4.1 ± 3.8 | 3.6 ± 3.6 | 2.3 ± 2.7 | 1.9 ± 2.4 | 3.3 ± 3.4 |
| GAD-7 score | 5.8 ± 5.0 | 5.0 ± 4.7 | 4.4 ± 4.8 | 2.7 ± 3.6 | 1.9 ± 2.9 | 4.0 ± 4.5 |
| FS20 functional status | 4.6 ± 2.1 | 4.8 ± 2.4 | 5.3 ± 2.6 | 4.9 ± 2.2 | 4.7 ± 2.0 | 4.9 ± 2.3 |

### Apparatus

All cognitive testing, including the figure drawing task, was administered on a Microsoft Surface tablet running the CCAB software platform. Drawing input was captured by direct fingertip contact with the capacitive touchscreen: the use of fingertip rather than stylus input was a deliberate design choice to facilitate at-home testing. Touch position was adaptively sampled at 30–60 Hz and recorded as time-stamped (x, y, t) coordinates with millisecond resolution. Finger-up and finger-down events were registered at the start and end of each contiguous fingertip contact, defining strokes as a single continuous trajectory between a touch and lift. The drawing canvas displayed on the tablet was a blank white rectangular field with fixed aspect ratio matched to the rendered template space.

### Figure copy and recall task

The CCAB figure drawing copy and recall test is modeled on the BCFT (Possin et al., 2011; Kramer, 2011), with moderate figure complexity designed to balance scoring tractability against sensitivity to organizational and memory deficits. The target figure consists of twelve canonical elements arranged within a bounding rectangle. Each element is a discrete geometric object with a canonical position, size, and shape on a fixed template. Figure 1 (below) shows the target template alongside a representative drawing, with stroke color reflect latency.

**Figure 1.**
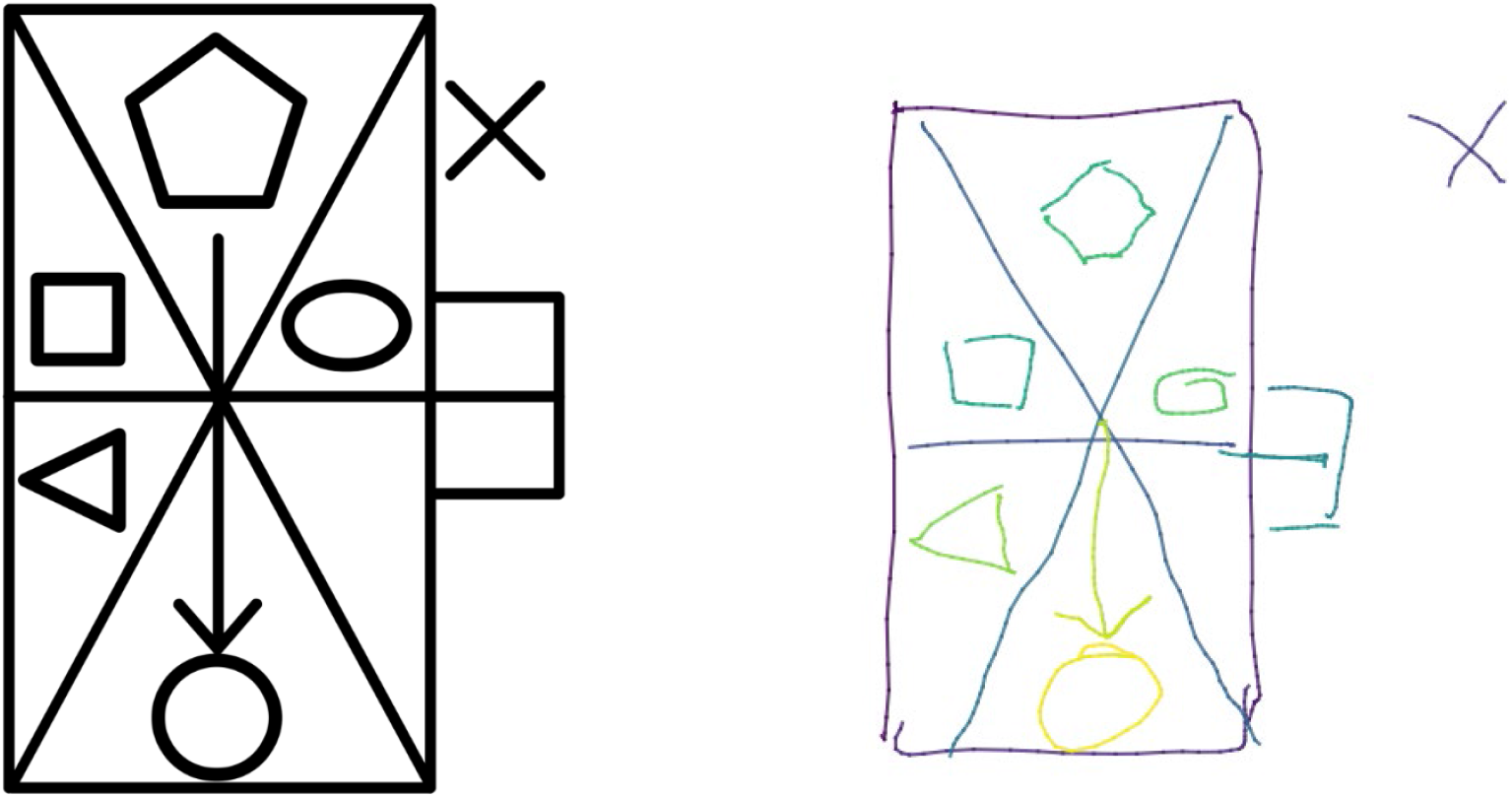
LEFT. The CCAB figure drawing target template. The figure consists of twelve canonical elements arranged within a bounding rectangle: the outer rectangular box (LR); two crossing diagonals forming a central X (LX); a pentagon in the upper region (PN); a small square on the mid-left (SS); an oval to the right of center (OV); a horizontal midline bisecting the rectangle (HL); a triangle in the lower-left quadrant (TR); a vertical shaft (VS) terminating in an arrowhead (AH); a circle at the lower portion of the figure (CO); an attached rectangle extending from the right edge (EB); and a small X positioned outside the main bounding rectangle on the upper right (SX). Each element occupies a fixed canonical position, size, and shape and was annotated as a discrete object with a bounding box for use in training the Vertex AI object-detection model described below. **RIGHT**. Drawing from one subject in the Copy condition. Line color codes latency.

The task was administered in two trials. During the Copy trial, the target figure was displayed on the left portion of the tablet screen, and the participant reproduced it onto a blank canvas in the right portion, with a 75 second time limit. The stimulus remained continuously visible throughout the trial. After approximately thirty minutes of intervening cognitive testing, the Recall trial was administered: the participant was asked, without prior warning at the time of the original copy, to reproduce the figure from memory on a blank canvas. Each trial produces a complete record of fingertip position at an adaptive sampling rate of 30 to 60 Hz.

### Image rendering and template registration

Completed drawings were rasterized from the x-y position log to a fixed-resolution PNG image and aligned to canonical 512 × 512 pixel template space. Registration was performed by detecting the four corners of the participant’s outer bounding rectangle using an expanding-search algorithm that extended search bands inward from each canvas edge until the longest contiguous horizontal and vertical line segment were identified. The detected corners were coregistered to the four canonical corner coordinates of the template and the figure was homographically aligned onto the template coordinate system. For drawings in which the outer rectangle was absent, fragmented, or grossly distorted (< 2% of cases), affine registration based on the centroid and principal axes of the drawing were used.

### Model architecture and training

Element detection was performed using a convolutional object detection model trained on the Vertex AI AutoML Vision platform (Google Cloud). The base architecture was a pretrained CNN producing bounding-box predictions with associated class labels and confidence scores. Early convolutional layers extract local features (edges, corners, line terminations); middle layers compose these into mid-level structural primitives (L-shapes, X-junctions, parallel lines, closed contours); deep layers combine these primitives into element-level templates corresponding to the twelve canonical figure elements.

The model was fine-tuned on a domain-specific corpus of 1,774 figure drawings drawn from the entire CCAB dataset (9,117 drawings, including those from longitudinal testing), randomly sampled across both trial types (Copy and Recall) so that the training distribution reflected the full range of element appearances that the deployed model would encounter. Ground-truth element labels and bounding boxes for the training corpus were annotated by a single expert rater (DLW) who applied fixed scoring rules across all 1,774 drawings to ensure consistency at the labeling stage. The corpus was partitioned into training, validation, and test splits at the image level (with all trials from a given subject assigned to the same split to prevent leakage).

Training optimized intersection-over-union (IoU) loss between predicted and ground-truth bounding boxes, with cross-entropy loss for class label assignment. The model produced a set of candidate detections per input image, each consisting of an element label (one of twelve element types), a bounding box (specified in 512 × 512 template coordinates), and a confidence score in the range of 0.0 -1.0. Non-maximum suppression was applied within each class to remove overlapping duplicate detections. Confidence calibration was performed for each element class: the confidence threshold maximizing hits vs. false alarms against ground-truth labels yielded element-specific thresholds ranging from 0.32 to 0.74. These element-specific thresholds reflected the differing element distinctiveness: elements with broad, recognizable templates (LR, CO) were reliably detected at high confidence, while elements resembling fragments of larger structures (TR, SS) required lower thresholds. For two elements (VS and HL) bounding-box expansions were applied to the detected boxes prior to scoring to compensate for the model’s tendency to draw tight boxes around line elements.

### Element scoring

Elements were scored along two dimensions: presence (whether the element was detected) and location (the distance of the element’s bounding box to that of the corresponding template element).

#### Presence

For each canonical element, the element was scored as present if a drawing component exceeded the element-specific confidence threshold. Presence was a binary indicator (0 or 1), summed across the twelve elements to yield the Element score, ranging from 0 to 12 and corresponding to the standard “elements present” count in manual scoring.

#### Location

For each element detected, Location was scored as 1.0 if the center of mass (COM) of the detected element’s bounding box fell within the bounding box of the corresponding template element, 0.5 if there was overlap between the two bounding boxes, and 0.0 otherwise. The Location score was the sum of per-element location scores across elements present in the drawing, ranging from 0 to 12.

The trial-level Total score combined presence and location into a composite score ranging from 0 to 24, with each element contributing one point for presence and up to one point for location, mirroring the per-element ELE+LOC structure of manual scoring. The three trial-level scores—Element, Location, and Total—were computed independently for the Copy and Recall trials, yielding six primary outcome scores per participant. In addition, a Memory Cost score (Recall − Copy), expressed in z-score units. Additional element-level metrics related to perseveration (duplicate detections), organization (drawing order), and shape distortion were computed and stored alongside primary scores.

### Vertex AI comparison with manual scoring

To evaluate inter-rater reliability and to validate the automated pipeline against expert human scoring, a random subsample of 1300 drawings was independently scored by three expert human raters using the twelve-element rubric described above. These drawings were drawn at random from the full corpus of 9,117 drawings across all testing waves. Each rater scored 400 unique drawings for comparison with Vertex AI, with an additional 100 drawings (52 Copy, 48 Recall) scored by all three human raters and the Vertex pipeline. Inter-rater analyses were conducted under a dual-sample structure: human–human pairwise correlations were computed on the 100-drawing with three-way overlap, while Vertex–human pairwise correlations were computed on each rater’s full set of 500 drawings. Raters scored independently and were blinded to one another’s scores. We also report agreement among all four scorers (three humans plus the Vertex pipeline) on the matched 100-drawing subsample to compare automated scores to the human-rater consensus.

### Motor-temporal pipeline

In addition to the static-image scoring described above, a parallel motor-temporal analysis pipeline was developed to extract stroke-level timing and kinematic features from the time-stamped position log from each drawing, including drawing time, pause time, intersegment pauses, and drawing velocity.

### Statistical analysis

Four sets of analyses are reported in the present manuscript. First, to evaluate inter-rater reliability and compare the automated pipeline within the human-rater consensus, three agreement metrics were computed, corresponding to the conventional inter-rater reliability measure used in the figure-drawing literature. Element-level (ELE) and Location-level (LOC) total-score correlations were computed to assess whether agreement on element presence and location. Per-element percent agreement was also computed for each of the twelve elements separately on the three-way overlap.

Second, to characterize demographic influences on automated scores, Pearson correlations were computed between each trial-level Vertex score (Element, Location, Total, separately for Copy and Recall) and a panel of demographic and clinical predictors including age, gender, years of education, vocabulary score, self-reported race/ethnicity, comorbidity count, daily medication count, the CFW, the GDS, the GAD-7 scale, daily computer use, daily reading hours, and the FS20 functional status measure. These analyses established whether the automated scores reproduced the demographic gradients (age decline, education and vocabulary advantage) expected from previously published studies.

Third, to characterize age-related decline on the figure drawing task, Copy total scores, Recall total scores, and Memory Cost scores (Recall − Copy) were converted to z-scores using the full-sample mean and standard deviations. Group means and standard errors were computed within each of age quintile. Pearson correlations between continuous age and z-score measure were also computed for the full sample.

Fourth, to characterize test–retest reliability and retest learning, paired Pearson correlations and paired-samples t-tests were computed separately for Copy and Recall trials. Analyses compare baseline scores and retest scores (using the same template drawing) at 1 day, 6 months, 18 months, and 30 months, on Total, Element, and Location scores, as well as motor and timing measures. Effect sizes for practice-related mean changes are reported as Cohen’s Δz based on the standard deviation of within-subject differences.

All correlations are reported with two-tailed p-values. Given the large sample size and the exploratory character of the analyses, we report nominal significance (p <.05,.01,.001) without familywise correction but emphasize effect-size magnitude over significance in the interpretation. Sample sizes vary slightly across analyses due to per-trial completion patterns and per-element missingness; exact N is reported for each correlation. Analyses were conducted in Python (pandas, numpy, scipy.stats) and R (base, psych).

## Results

### Inter-rater reliability and agreement of Vertex AI with human raters

The mean Pearson correlation among the three human raters on Total scores across all trials (computed on the 100-drawing overlap) was r = 0.972, while the mean Vertex–human vs. human correlation across the three rater corpora was r = 0.966, corresponding to a Vertex–human gap of approximately 0.006. Agreement was higher on Recall trials (human–human r = 0.965; Vertex–human r = 0.951) than on Copy trials (human–human r = 0.851; Vertex–human r = 0.803) for all comparisons, reflecting the wider score range on Recall (group mean Total ≈ 16 vs. ≈ 22 on Copy) which provides greater dynamic range for the correlation. The compressed Copy correlations are a known consequence of ceiling effects on Pearson r and are not specific to the Vertex pipeline.

Decomposing the Total score into Element (presence) and Location components revealed an interpretable structure. On the Element-only total, Vertex agreement with humans (mean r = 0.959) was nearly indistinguishable from the agreement among the human raters themselves (mean r = 0.963), indicating that the automated pipeline detected element presence as accurately as expert human raters.

However, small human vs. Vertex disagreement emerged in Location scores, particularly for smaller elements, as shown in Figure 2. Vertex agreement with humans on Location (mean r = 0.951) fell slightly below the human–human mean (r = 0.972). Bounding boxes were not visible to human raters so their ability to judge bounding box overlap and center-of-mass distances was imprecise. As a result, mean Total scores were systematically lower for the Vertex pipeline than for the human consensus (offsets −0.41 to −1.03 on Copy and −0.32 to −2.08 on Recall). This absolute calibration offset did not affect the rank-order agreement that Pearson correlations measure: Vertex ranks drawings consistently with human raters while assigning somewhat lower absolute location scores. The per-element percent exact agreement on the 100-drawing three-way overlap is summarized in Table 2.

**Table 2.** Per-element identification and percent exact agreement on the 100-drawing three-way overlap, for element presence (ELE) and location (LOC). One hundred drawings were scored by all three raters and the Vertex pipeline. The “Vertex count” column gives the number of drawings where the Vertex pipeline identified that element. Human columns measure agreement among the three expert raters: the all-3 column shows complete agreement, the pairwise column averages concordance over the three rater pairs. The two Vertex agreement columns give the percentage of drawings on which the automated score matched the human majority consensus for presence and location. Elements are ordered by decreasing all-3 human agreement.

| Element | Name | Vertex count | ELE all-3 (%) | ELE pairwise (%) | LOC all-3 (%) | LOC pairwise (%) | Vertex ELE match (%) | Vertex LOC match (%) |
| --- | --- | --- | --- | --- | --- | --- | --- | --- |
| LR | large rectangle | 99 | 99.0 | 99.3 | 99.0 | 99.3 | 100.0 | 100.0 |
| LX | large X (diagonals) | 94 | 98.0 | 98.7 | 100.0 | 100.0 | 97.0 | 96.8 |
| HL | horizontal line | 95 | 97.0 | 98.0 | 90.0 | 93.3 | 99.0 | 65.3 |
| CO | circle | 75 | 89.0 | 92.7 | 98.0 | 98.7 | 92.0 | 94.7 |
| PN | pentagon | 79 | 93.0 | 95.3 | 91.0 | 94.0 | 97.0 | 97.5 |
| EB | external box | 78 | 92.0 | 94.7 | 89.0 | 92.7 | 100.0 | 93.6 |
| AH | arrowhead | 84 | 92.0 | 94.7 | 88.0 | 92.0 | 96.0 | 73.8 |
| OV | oval | 87 | 90.0 | 93.3 | 90.0 | 93.3 | 93.0 | 90.8 |
| SS | small square | 74 | 91.0 | 94.0 | 87.0 | 91.3 | 95.0 | 89.2 |
| TR | triangle | 80 | 91.0 | 94.0 | 75.0 | 83.3 | 97.0 | 92.5 |
| VS | vertical shaft | 62 | 75.0 | 83.3 | 82.0 | 88.0 | 90.0 | 96.8 |
| SX | small x | 70 | 72.0 | 81.3 | 53.0 | 68.7 | 98.0 | 77.1 |
| Mean |  | — | 89.9 | 93.3 | 86.8 | 91.2 | 96.2 | 89.0 |

**Figure 2.**
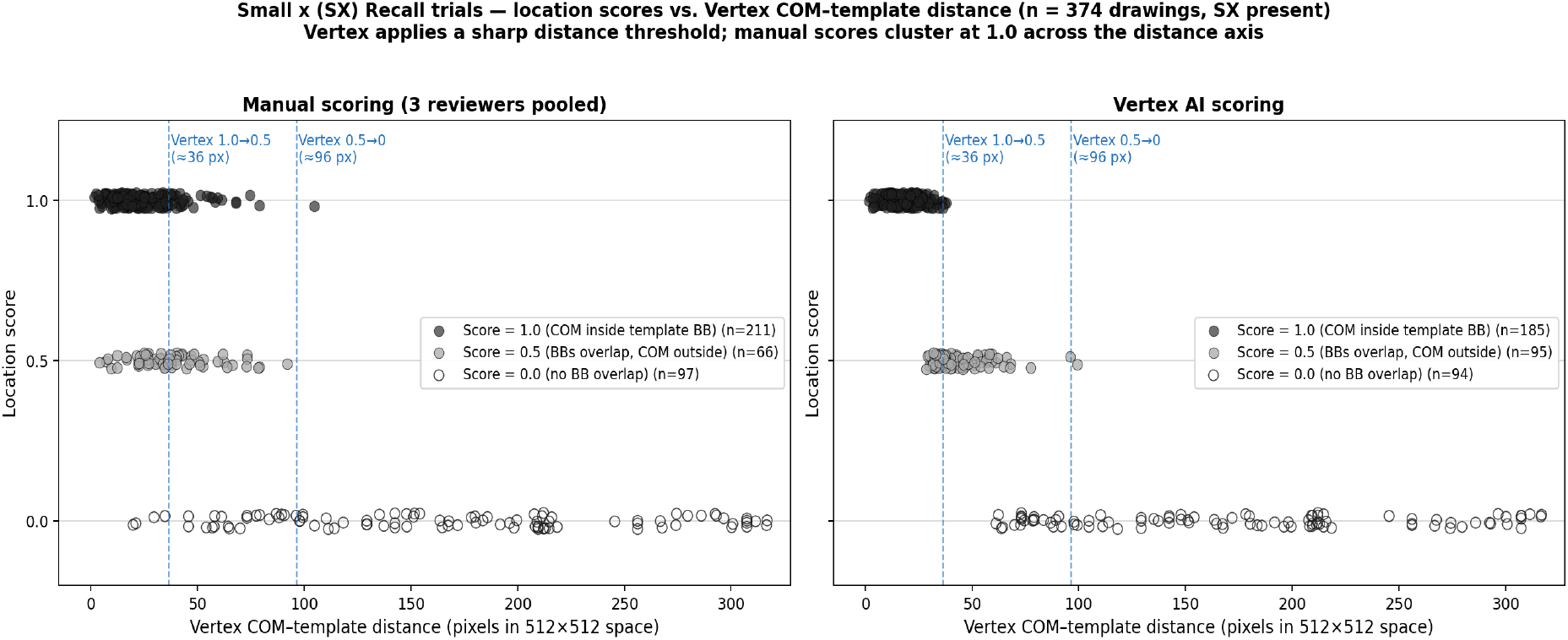
Comparison of manual (left) and Vertex AI (right) location scores for the SX element (small x outside the rectangle) on Recall trials, plotted as a function of the Vertex-measured distance in pixels between the center of mass (COM) of the SX bounding box and the SX template bounding box. Data from 374 drawings scored by manual reviewers (**LEFT**) and the Vertex pipeline (**RIGHT**). Manual scores showed imprecision due to the difficulty of estimating the COM and size of SX bounding boxes. Location scores of 1.0 require the center-of-mass (COM) of the element’s bounding box to fall within the bounding box of the corresponding template; scores of 0.5 require overlap between the two bounding boxes. Location scores of 0.0 (bottom right) can occur at small distances for tiny SX element drawings.

### Training bias

Although the Vertex model was trained on labels from a single expert (DLW), automated score agreed similarly for the three human raters: Vertex–DLW r = 0.969, Vertex–KH r = 0.972, and Vertex–IJ r = 0.956. The Vertex correlation for the rater whose labels trained the model (DLW) minus the mean of correlations of other two raters (KH and IJ) was +0.005, indicating no preferential advantage. Vertex agreement with the 2-of-3 majority consensus on the three-way overlap was r = 0.978, equaling the highest single human–human pair. Together these results indicate that the Vertex model captured generalizable scoring criteria rather than rater-specific biases, and that the training-data composition (single expert annotator) did not limit the model’s applicability across independent expert raters.

### Demographic and clinical correlates of figure drawing performance

Pearson correlations between figure drawing Total scores on for the Copy and Recall trials and the demographic and clinical predictor panel are summarized in Table 3. For race, a categorical variable, we report mean Total scores by group along with the omnibus one-way ANOVA.

**Table 3.** Demographic and clinical correlates of figure drawing Total scores at baseline. Pearson correlations between each continuous predictor and the Copy Total and Recall Total scores in subjects with both Copy and Recall drawings at baseline who also had questionnaire data (n = 2,011; per-predictor N is slightly lower for predictors with item-level missingness and is reported in the final column). Significance markers: * p <.05, ** p <.01, *** p <.001. Categorical race results are based on self-reported race.

| Predictor | r (Copy) | r (Recall) | N |
| --- | --- | --- | --- |
| Age (years) | −0.17*** | −0.32*** | 2,011 |
| Education | +0.19*** | +0.13*** | 2,010 |
| Vocabulary (CCAB QSIN) | +0.25*** | +0.13*** | 2,011 |
| Female (vs. male) | +0.05* | +0.04 | 1,980 |
| Daily medication count | −0.18*** | −0.23*** | 2,000 |
| Comorbidity count | −0.12*** | −0.14*** | 2,011 |
| Daily computer use (hrs) | +0.14*** | +0.18*** | 1,993 |
| Geriatric Depression Scale | −0.02 | +0.05* | 2,008 |
| Generalized Anxiety Disorder | −0.07** | −0.00 | 2,005 |
| Cognitive Failures Questionnaire | −0.05* | +0.01 | 2,009 |
| FS20 functional status | −0.10*** | −0.06** | 1,995 |
| Race (mean Total) |  |  |  |
| White | 22.49 | 14.42 | 737 |
| Black | 21.04 | 13.07 | 412 |
| Asian | 22.61 | 15.72 | 342 |
| Other | 22.09 | 14.89 | 518 |
| ANOVA Race | $F(3,2005) = 40.0, p < .001$ | $F(3,2005) = 22.5, p < .001$ | 2,009 |

Age was a significant negative predictor of performance, with a substantially larger influence on the Recall trial than (r = −0.32 (p <.001) than on of the Copy trial (r = −0.16). Education and vocabulary were also significant predictors with larger correlations on Copy than on Recall trials (education, r = +0.19 Copy vs. +0.13 Recall; vocabulary total r = +0.25 Copy vs. +0.13 Recall). The pattern is consistent with education and vocabulary indexing the cognitive reserve that support the planning, perceptual organization, and motor execution that the copy trial demands, with smaller incremental contribution to the memory-specific demands of the recall trial.

Daily medication count was a substantial negative predictor of both trials, with a slightly larger effect on Recall (r = −0.23) than on Copy (r = −0.18); the comorbidity count showed a similar pattern (Recall r = −0.14; Copy r = −0.12), with both correlations reaching the p <.001 threshold in the present sample. The medication and comorbidity correlations index general health burden and broadly affect figure drawing performance rather than dissociating cleanly between visuoconstruction and memory. Conversely, daily computer use was a positive predictor of both trials with a slight Recall advantage (Recall r = +0.18; Copy r = +0.14). Per-element analyses indicated that the race-related Total-score gap on the Copy trial was broadly distributed across elements rather than concentrated on any single element.

### Age-related decline

Figure drawing z-scores stratified by quintile are shown for Age in Figure 3A, and stratified by vocabulary quintile in Figure 3B, with Copy trial (sub-panel A), Recall (sub-panel B), and a Memory Cost score (sub-panel C, Recall − Copy in z units.

**Figure 3.**
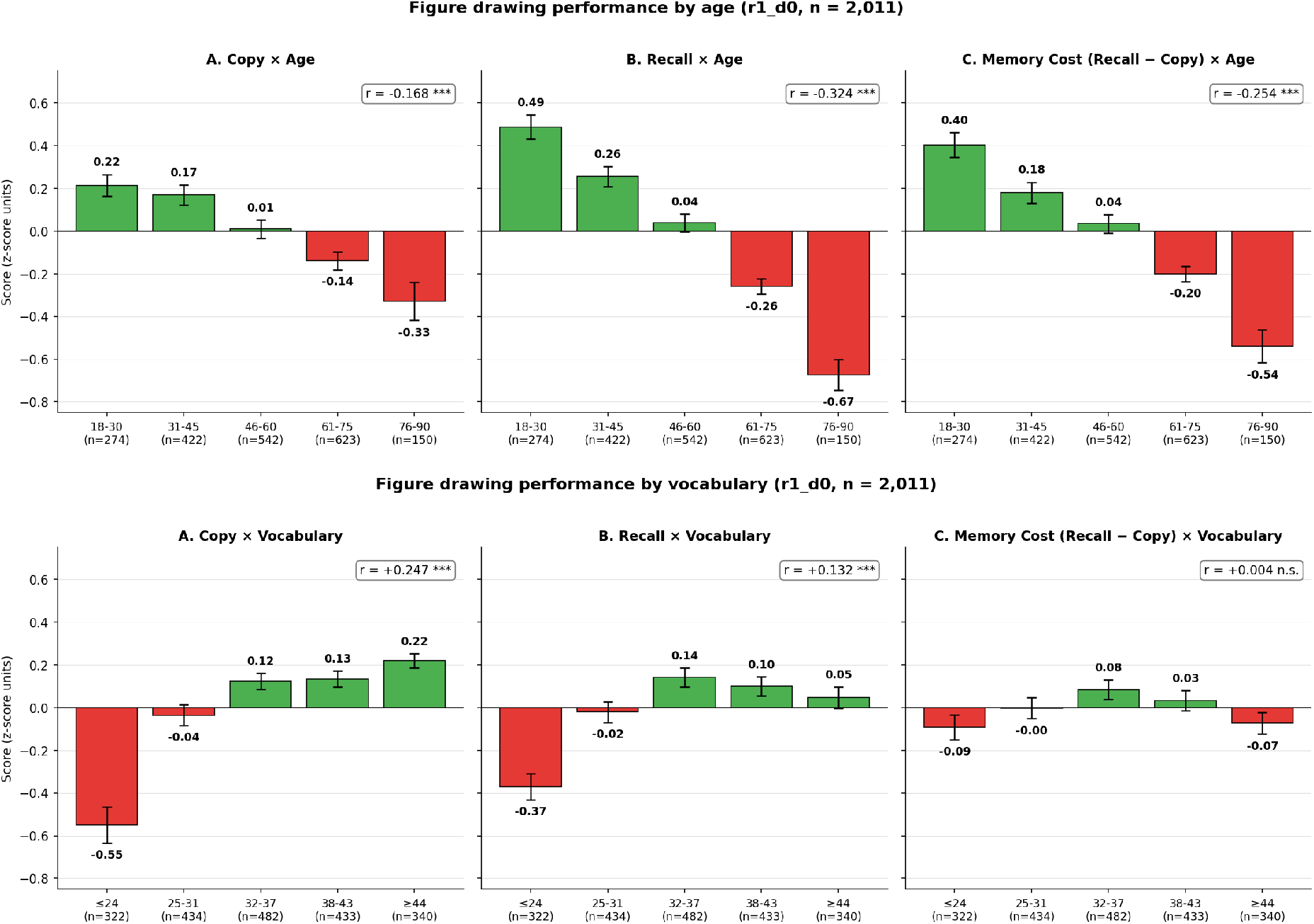
Figure drawing performance by age and vocabulary at baseline. **(A)** Top row: mean Copy total (left), Recall total (center), and Memory Cost (Recall − Copy; right) z-scores, stratified by five age quintile (n = 2,011). **(B)** Bottom row: the same three measures stratified by five vocabulary quintile based on the CCAB vocabulary score. In both rows all sub-panels share a common z-score y-axis, error bars indicate the standard error of the mean, and continuous Pearson correlations between the stratifying variable and each z-scored measure are shown in the upper-right of each sub-panel.

Signifcant age effects were seen. First, all measures showed a monotonic age-related decline that survived at the p <.001 threshold. Second, the magnitude of the age-related decline differed substantially for different measures. The Copy gradient was the shallowest, with a continuous correlation of r = −0.17; the Recall gradient was the steepest, with a continuous correlation of r = −0.32, approximately twice the magnitude of the Copy correlation. The Memory Cost gradient fell between the two (r = −0.25), with quintile means spanning from +0.40 z to −0.54 z. The persistence of a clear age gradient on the Memory Cost score, after the participant’s own copy performance is subtracted, demonstrates that age-related decline on the Recall trial is not merely a downstream consequence of declining visuoconstructional ability. Rather, it reflects an additional, memory-specific component captured by the recall trial.

Vocabulary showed a different pattern from age. Vocabulary predicted both trials positively, somewhat more strongly for the Copy trial (r = 0.25) than the Recall trial (r = 0.13), rising monotonically across quintiles from −0.55 z in the lowest-vocabulary quintile to +0.22 z in the highest on Copy and from −0.37 z to +0.05 z on Recall. Because vocabulary raised Copy and Recall in parallel, it had essentially no relationship with the Memory Cost score (r = 0.004, n.s.). This contrasts sharply with age, which drove a strong Memory Cost gradient (r = −0.25). Thus, higher vocabulary is associated with better figure drawing overall—consistent with vocabulary indexing crystallized cognitive reserve—but, unlike age, does not selectively influence the memory-dependent recall trial relative to copy.

### Figure organization and duplicates

In addition to element-level presence and location, the Vertex pipeline produces figure-level qualitative measures derived from element detections, including an organization score (0–1) indexing the large-to-small drawing strategy, large- and small-element counts, and a duplicate count (n_dupes, the number of supernumerary or mislocated element detections). Two features of these measures were of note. The duplicate count was approximately eight-fold higher on Recall than on Copy (mean 0.47 vs. 0.06; paired-samples t for the condition difference p < 10^−100^), with roughly 47% of healthy subjects producing at least one duplicate on Recall versus only 6% on Copy This reflected the well-known tendency to produce extraneous, perseverative elements when reconstructing the figure from memory (Sedda et al., 2012; Pelati et al., 2011). The small-element count on Recall declined with age (r = −0.28), consistent with the established finding that figure details are the first features to drop out of complex-figure recall with age (Machulda et al., 2007).

### Test–retest reliability and retest learning

Subsets of the normative sample repeated the figure drawing task at 1-day (overnight) retest and at 6-, 18-, and 30-month follow-ups. The overnight retest provides the cleanest estimate of measurement stability, minimally affected by underlying cognitive change. Total-score test–retest reliability at 1 day was higher on the Recall trial (r = 0.72, n = 972) than on the Copy trial (r = 0.49, n = 974), and remained substantial across the longer intervals (Recall r = 0.61 at 6 months, 0.62 at 18 months, and 0.55 at 30 months). The lower Copy reliability reflected substantial Copy ceiling effects (25.3% of Copy drawings scores were at the 24-point ceiling), which left little dynamic range for the Pearson correlation.

A large overnight learning effect was evident on the Recall trial: mean Recall Total scores rose from 14.0 to 18.1 (of 24) from baseline to the next day (paired t = 36.7, p < 10^−180^, Δz = +1.18), whereas the Copy trial was essentially unchanged (Δz = +0.18). Crucially, this Recall gain did not fully decay but cumulated with repeated exposure: relative to baseline, Recall Total scores remained elevated at 6 months (Δz = +0.34), 18 months (Δz = +0.50), and 30 months (Δz = +0.75), indicating prior exposure benefit that increased over two and a half years (Figure 4).

**Figure 4.**
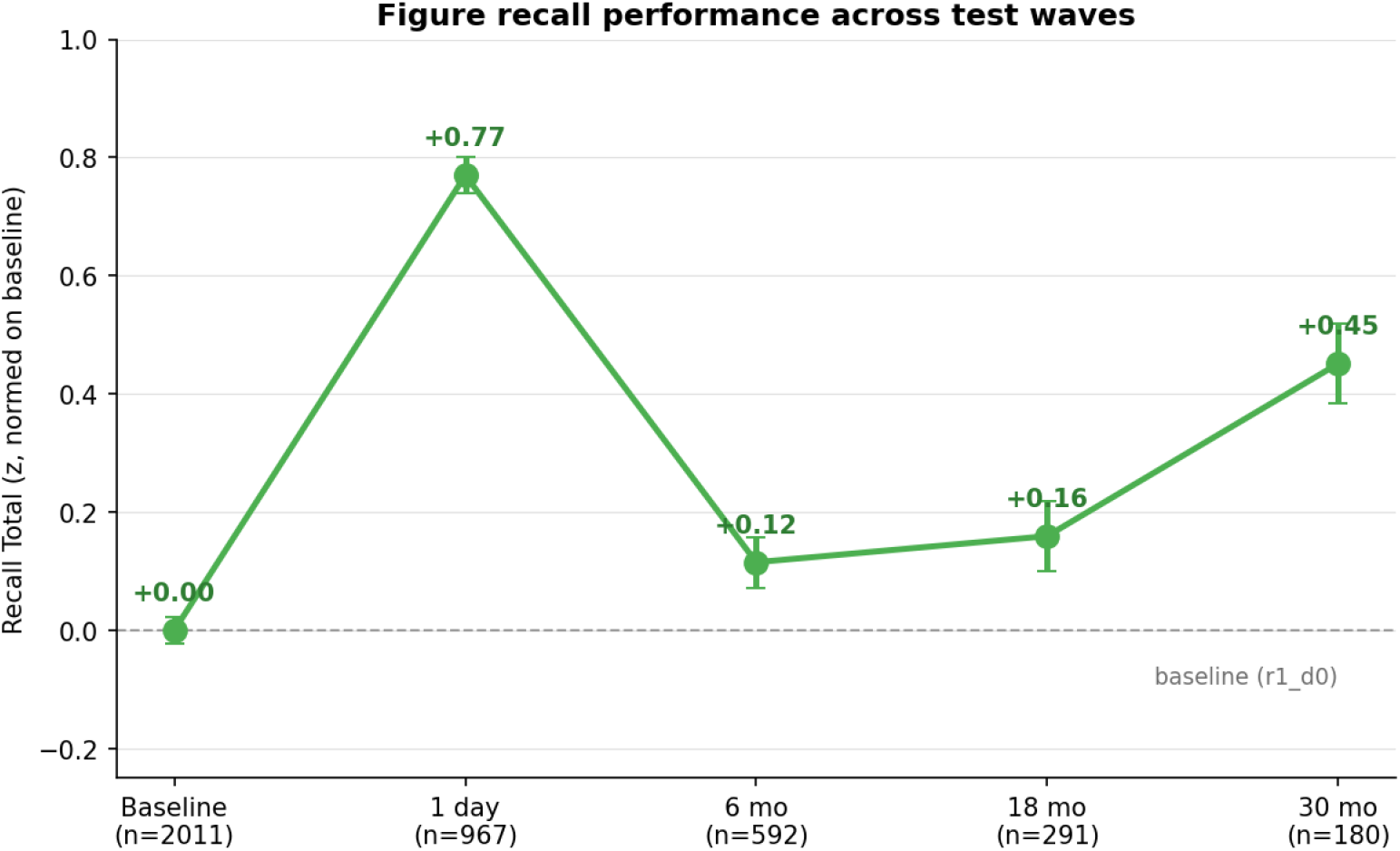
Figure recall performance across test waves. Mean Recall Total score at each wave in the analytic sample (n = 2,011), expressed in z units normed on the baseline (r1_d0) Recall distribution (mean 14.5 of 24, SD 4.67); the dashed line marks the baseline mean. Recall peaked at the 1-day retest (+0.77 z), declined by 6 months (+0.12 z), and remained above baseline at 18 (+0.16 z) and 30 months (+0.45 z); error bars indicate the standard error of the mean. Per-wave n varied with enrollment timing (and minimally attrition) and is shown below each point. Note that these z scores were normed on the baseline score distribution and therefore differ from the within-subject paired effect sizes reported in the text (e.g., the overnight Δz = +1.18) which were normed on the SD of the change scores).

### Motor and timing measures

Trial-level timing and segment-count metrics from the motor-temporal pipeline complemented the static element-level scores. Several patterns emerged. First, on Copy trials both completion time (41.8 ± 13.4 s) and drawing time (21.7 ± 7.7 s) were essentially uncorrelated with age, indicating that healthy adults across the full age range produce the figure at a comparable rate with the stimulus in view. On Recall, drawing time was likewise uncorrelated with age but mean inter-stroke pauses (3.70 ± 2.62 s) lengthened substantially with age (r = 0.19), showing that the older-adult slowdown on Recall reflects time spent retrieving and planning between strokes rather than slower stroke execution. Second, the number of strokes decreased with age on Recall (r = −0.13) but not Copy, consistent with element omissions on the Recall trial. Third, the within-stroke speed coefficient of variation (0.58 ± 0.11 Copy, 0.57 ± 0.11 Recall) showed an age effect on both trials (r = 0.17 Copy, 0.11 Recall), with older adults producing jerkier strokes even when the figure was in view. The timing and motor data therefore provide a process-level account of performance that complements the static element-level Memory Cost: when the figure is absent and the participant draws from memory, older adults pause longer, omit more elements, and draw less smoothly than they do on the same figure with the template visible.

The six trial-level motor and timing measures showed overnight reliabilities similar or slightly lower than those of Recall scores (drawing time, Copy 0.76 / Recall 0.73; stroke speed 0.65 / 0.69; stroke count 0.62 / 0.62; speed CV 0.46 / 0.53; total pause time 0.57 / 0.41; mean inter-segment pause 0.43 / 0.34), and changed only modestly across the 6- to 30-month intervals.

## Discussion

Here, we describe a computer vision pipeline for scoring the figure copy and recall tests of the CCAB. The Vertex AI pipeline produced Element scores that were indistinguishable from those of human raters, and that produce Location scores that were slightly more precise —and modestly different —from the human consensus. The observed Vertex–human gap with human scores (∼ 0.006) was approximately one-tenth the size of the corresponding gap reported by Vogt et al. (2019), the most direct comparison in the existing literature. The element-level decomposition we adopted contrasts with the global image-classification approach used by Vogt et al., and other end-to-end deep-learning systems that collapse the spatial structure of the drawing into a fixed-length feature vector producing a single score. The Vertex object-detection model instead produces element and location scores directly comparable to those produced in manual scoring.

Other recent end-to-end approaches (Park et al., 2023; Langer et al., 2024) have reported strong absolute-error performance using CNNs trained on much larger corpora (more than 20,000 drawings). In contrast, our pipeline used a smaller training corpus (1,774 drawings, with ∼20,000 elements) and trained the model to detect each canonical element independently rather than to predict a global score.

The decomposition of inter-rater agreement into Element and Location components clarifies what drives the small Vertex–human scoring gap. Element identification was essentially identical between automated and manual scoring, while the small Location gap reflected improved Vertex precision in applying the center-of-mass scoring rules with single-pixel precision, which was impossible for human reviewers lacking visible bounding boxes.

A further advantage of the element-level architecture is that it yields reliable scores for individual elements, not merely aggregate totals. This addresses a long-standing reliability challenge with manual figure-drawing scoring of individual figure elements: in the Tupler et al. (1995) study individual Rey-O item agreement ranged from 0.14 to 0.96. Because the present pipeline applies a deterministic, precise rule to every element, and per-element agreement with human raters was uniformly high (Table 2), Vertex scores are accurate enough to support qualitative error-pattern and process-based analyses without human intervention.

### Age-related decline and the Memory Cost dissociation

The age-related findings on the figure drawing task are consistent with a substantial literature documenting age-related decline in visuoconstruction and visual episodic memory. Recall total scores declined with age at approximately twice the rate of Copy total scores, and the Memory Cost score showed a similar decline. The persistent age gradient after Copy performance is subtracted demonstrates that age-related decline on the recall trial is not merely a downstream consequence of declining visuoconstructional ability but reflects an additional, memory-specific component. This is precisely the clinical signal that motivates the inclusion of a recall trial in figure-drawing testing and that copy-only screening tests, such as the Clock Drawing Test and the interlocking pentagons of the MMSE, cannot capture.

### Retest learning

The overnight retest revealed an exceptionally large learning effect on the Recall trial (Δz = +1.18) following one prior exposure to the figure. The Recall gain persisted and grew across repeated presentations at longer follow-ups, remaining well above baseline at 30 months (Δz = +0.75). Individual differences in Recall remained highly stable across all intervals (r = 0.55–0.72) indicating that the recall trial reflects a durable memory representation.

### Advantages and validity of the automated scoring pipeline

Automated scoring of the CCAB figure drawing task offers four concrete advantages over expert manual scoring. First, the pipeline requires no additional time from the examiner: drawings are scored automatically without inter-rater variability or drift. Second, the pipeline is perfectly replicable: the same drawing scored twice by the same pipeline produces identical scores, without intra-rater drift or disagreement among reviewers. Third, the pipeline applies the location-scoring rubric with single-pixel precision that expert human raters cannot match. Fourth, the pipeline provides reliable motor and timing measures that manual scoring cannot recover.

Beyond these practical advantages, automated scores captured the cognitive constructs the test is designed to measure. The automated scores recovered expected demographic and clinical correlates in our normative sample, including the approximately twofold larger age correlation on the Recall trial driving an age-related Memory Cost gradient (r = −0.25). In contrast, while vocabulary raised Copy and Recall in parallel and was unrelated to Memory Cost (r = 0.004, n.s.). Together with the inter-rater equivalence on element identification reported above, these lines of evidence establish that the automated pipeline produces scores that are valid indices of the cognitive constructs the test is designed to measure rather than figure-specific image features that the object-detection model has learned to weight idiosyncratically.

### Limitations and future directions

The motor-temporal analyses reported here are restricted to trial-aggregate timing, segment, and motor metrics that are robust at sampling rates of 30 Hz and above, and the analyses excluded drawings sampled below 30 Hz from the tablet’s adaptive-sampling distribution. Both the CCAB tablet and similar capacitive-touchscreen platforms sample adaptively rather than at a fixed rate, and roughly 1% of drawings fall below 30 Hz; these drawings are excluded from the motor analyses because kinematic features are unreliable at very low sampling rates. Trial-level ratios and aggregate counts — such as completion time, drawing time, pause time, segment count, total path length, mean stroke speed, and within-stroke speed coefficient of variation — are constructed from quantities that integrate over hundreds of samples per drawing and remain reliable across the 30–60 Hz range.

The present model is figure-specific. Although the approach is generalizable, each new figure would require its own training corpus.

## Conclusion

Automated scoring of figure-drawing tests has historically been difficult because the construct is multidimensional, the scoring rules are intricate, and the agreement structure among expert human raters is itself imperfect, particularly at the item level. The Vertex AI pipeline described here addresses these difficulties by decomposing the scoring problem into element-level presence and location judgments that mirror expert manual scoring, by training an object-detection that provides element level scores similar to those produce by expert raters rather than collapsing them into an opaque global score. The pipeline agrees with expert raters at essentially the same level they agree with one another, recovers the expected demographic and clinical correlates of figure drawing performance, reveals a memory-specific age-related decline, and exhibits test– retest reliability comparable to other long-form cognitive measures. Its current deployment in the CCAB platform removes the principal practical barrier — reviewer burden and uncertainty — that has limited the use of figure copy and recall tests in routine clinical neuropsychological practice.

## Declarations

## Funding

This work was supported by the National Institute on Aging (NIA R44AG097322). The funder had no role in study design, data collection and analysis, decision to publish, or preparation of the manuscript.

## Competing interests

All authors are employees of Neurobehavioral Systems, Inc.

## Ethics approval and informed consent

All subjects completed informed consent under a protocol approved by Western Institutional Review Board (WIRB protocol 20201196).

## Clinical trial registration

Clinical trial: NCT04800588 (https://clinicaltrials.gov/study/NCT04800588).

## Data availability

De-identified data underlying the analyses reported in this manuscript are available from the corresponding author on reasonable request.

## Author contributions

All authors contributed to the design, data collection, analysis, or writing of the present manuscript. All authors have read and approved the final version.

## Notes

Supported by NIA R44AG097322.

